# Community-based Case Studies of Vaccine Hesitancy and the COVID-19 Response in South Africa; The VaxScenes Study

**DOI:** 10.1101/2022.02.21.22271272

**Authors:** Charles Shey Wiysonge, Nancy Coulson, Nirvana Pillay, Sara Cooper, Candice Groenewald, Zaynab Essack, Saahier Parker, Gregory Houston, Jane Simmonds, Anelisa Jaca, Muyunda Mutemwa, Patrick DMC Katoto, Heidi van Rooyen

## Abstract

**Background:** South Africa has reported more than half of all COVID-19 cases and deaths in Africa. The South African government has launched a large COVID-19 immunization campaign with the goal of reaching more than 40 million individuals. Nonetheless, certain international, largely internet-based surveys have shown a significant proportion of vaccine hesitancy in South Africa. This study aims to determine and co-create with local stakeholders a comprehensive understanding of vaccine hesitancy and opportunities to support the promotion of other COVID-19 health-promoting behaviours at community level.

**Methods and design:** A mixed-methods, multiple case-study design; informed by the socio-ecological model of behaviour change. Four socio-economically diverse communities across South Africa will be selected and data collection will take place concurrently through three iterative phases. Phase 1 will provide insights into community experiences of COVID-19 (response) through desktop mapping exercises, observations, in-depth interviews, and focus group discussions (FGDs) designed as expression sessions with local stakeholders. Phase 2 will explore the extent and drivers of community acceptance of COVID-19 vaccines. This phase will comprise a quantitative survey based on WHO’s Behavioural and Social Drivers of Vaccination tool as well as further FGDs with community members. Phase 3 will involve cross-case study syntheses and presentation of findings to national role-players.

**Discussion:** This study will provide ground up, locally responsive, and timeous evidence on the factors influencing COVID-19 health-seeking behaviours to inform ongoing management and mitigation of COVID-19 in South Africa. It will also provide insights into the applicability of a novel vaccine hesitancy model in Africa.

## 1. Introduction

South Africa first went into national lockdown on 26 March 2020 in response to the coronavirus disease 2019 (COVID-19) pandemic and has since aligned with international guidance led by the World Health Organization (WHO) on how to manage the unprecedented pandemic in the face of emergent scientific evidence [1, 2]. Lockdown regulations implemented in varying degrees by the South African government responded to the waves of COVID-19 amidst increasing uncertainty and social and structural damage to South Africa [3]. During the first quarter of 2021, the country experienced serious levels of COVID-19 infections as the second wave of the COVID-19 epidemic unfolded across the country [4]. Internationally and locally, new variants of COVID-19 were found to be more infectious and the surge in COVID-19 cases at the beginning of 2021 was more serious than the infection levels experienced at the start of the pandemic in 2020 [5].

The scientific pursuit of biomedical interventions to manage COVID-19 saw the proliferation of studies on drug and vaccine efficacy during 2020 [6]. Over the year, the need to manage and translate the changing landscape of COVID-19 evidence as it emerged was ever-present. This complicated the implementation and maintenance of behavioural interventions regarding non-pharmaceutical interventions (NPIs), but also emphasised the need for NPIs given we had nothing else at the time. In early 2021, NPIs were the only available and accessible methods to reduce and prevent the spread of COVID-19 in the country [1, 2]. At the same time, the safety and efficacy of a number of COVID-19 vaccines had been established and the manufacture, procurement, and rollout of these vaccines was a global priority [7]. Contrary to high-income countries, at the beginning of 2021, limited numbers of vaccine doses had been procured for South Africa and ongoing procurement and rollout was uncertain [8]. However, vaccinating the South African population at scale with efficiency was a top government priority. COVID-19 vaccine uptake will involve behaviour change at a large scale as the government aims for a vaccination coverage of at least 67% of the South African population in order to control COVID-19 infections in the country [9].

Vaccine hesitancy is one of the main challenges to be addressed for the vaccine programme to work in South Africa [10]. Vaccine hesitancy, according to WHO’s global working group on “Measuring Behavioural and Social Drivers of Vaccination” (BeSD) is a “motivational state of being conflicted about, or opposed to, getting vaccinated” which includes intentions [11]. Limited evidence available in early 2021 showed wide variation in COVID-19 vaccine hesitancy from 18% to 48% across various time points and geographical locations in South Africa [12, 13]. Many elements could have contributed to the vaccine hesitancy. For example, previous experiences that communities have of the delivery of healthcare services could influence both vaccine intentions and uptake [14]. Contextual factors that shape the enabling environment for vaccine uptake within a middle-income country like South Africa might be expected to play a bigger role in levels of vaccine hesitancy when compared with the results found in other studies which are largely conducted in high-income countries [15, 16]. For example, the availability and cost of transport is regularly a barrier to access to healthcare services in resource constrained communities [17, 18]. In addition, the dominance of social media has been shown in a global study to directly impact on vaccine uptake [19]. Even before the COVID-19 pandemic, South Africans were increasingly finding themselves in echo chambers of fake information about vaccination on social media and the internet [20-26].

Ensuring optimal uptake of COVID-19 vaccines in South Africa would thus involve multiple factors including knowledge, creating an enabling environment, addressing social influences, and personal motivation [18, 27, 28]. The preceding sections highlight the need for a ground up, contextually informed approach, that recognises the lived experience and understanding of individuals and places them in the social world in which they live and work. That was the rationale for the proposed VaxScenes Study, which aims to determine and co-create with local stakeholders in four diverse South African communities, a comprehensive understanding of vaccine hesitancy and opportunities to support the promotion of COVID-19 health seeking behaviours.

## 2. Materials and Methods

The specific objectives are to:

1. Create a community-based COVID-19 relevant geographic information system (GIS) picture that can be used for local planning in the COVID-19 response, including for vaccination and NPIs to curb the spread of COVID-19.
2. Document stories of the lives of community members affected by COVID-19 sickness, death, and economic loss in four communities.
3. Use the socio-ecological model to provide a community level assessment of the barriers and enabling factors at various levels (personal, interpersonal, community, and social/political) to vaccine acceptance and for ongoing adherence to behavioural measures.
4. Pilot the BeSD survey tool to provide guidance on its application in South Africa and to baseline vaccine hesitancy at the local level.
5. Co-create with community-level stakeholders, context sensitive strategies to address vaccine hesitancy and COVID-19 health seeking behaviours to guide both local and national responses.

Four community-based case studies will be undertaken using the socio-ecological model [29]. The research methods to be applied in the case studies are described in further detail in the following sections. Each component of data collection will be complemented by a process of engagement with stakeholders at the community level, described as action research [30]. These stakeholders will be identified during community-level mapping, and will include local community leadership and service providers, such as health facility managers and school principals. These and other key informants will be interviewed and then invited to participate in workshops to contribute to the interpretation of findings and to co-create community-level strategies to address vaccine hesitancy and to strengthen the COVID-19 response as findings become available.

### 2.1. Conceptual framework

We propose using the socio-ecological model [29] for behaviour change as a framework to understand how to increase COVID-19 vaccine uptake by looking at barriers and enabling factors at various levels, ranging from the individual to the interpersonal (the influence of family, peers) to the community, and to the wider context (policy, infrastructure, resources). Health messaging and correct information, although important, is often not enough for people to implement sustainable behaviour change. The socio-ecological model acknowledges that for any behaviour change to be maintained and sustained, it needs to happen at many different levels. Information about the COVID-19 vaccine in isolation to other interventions will not be sufficient to get the uptake required for herd immunity; information and strategies that address vaccine hesitancy and that motivate people to access COVID-19 vaccines are also needed. It is critical to behaviour change in a community that all four of the levels - personal, interpersonal, community, and social/political - be capacitated for a change in behaviour to occur in many people and then be maintained and sustained. Box 1 illustrates how the different levels of the socio-ecological model can be applied to address vaccine hesitancy. In practice, we will contextualise tools recently developed by the BeSD working group and use them in this study. The BeSD framework consists of four domains that could potentially shape vaccine intentions and uptake, namely, what people think and feel about vaccines; social processes that drive or inhibit vaccination; individual motivations (or hesitancy) to seek vaccination; and practical factors that shape the experience of seeking and receiving vaccination [14].

#### Box 1

**Applying the socio-ecological model to address vaccine hesitancy**

*<u>Individual (Self)</u>*: understand the information about the vaccine (know that it is available and it is recommen you get it), be motivated to get it, and be able to get access

*<u>Interpersonal</u>*: getting the vaccine needs support from partners, family members, and peers, or at least encour for this behaviour

*<u>Community</u>*: getting the vaccine needs to become a social norm and the socially accepted and expected thi You need to be able to access the vaccine easily and safely at a local primary healthcare facility, doctor or wo Religious organisations, community organisations, local leaders, traditional leaders, traditional healers, a influencers (social and political) need to be supportive of vaccine uptake

*<u>Environment</u>*: At a public policy level, the availability of the vaccine needs to be secured and the vaccine ne available. Resources need to be available to place vaccination centres within easy reach of the community.

The BeSD tools include qualitative tools (in form of guides for in-depth interviews with stakeholders), quantitative tools (that is, questionnaires for population surveys), and a guidebook to support implementation of the tools. The tools focus on childhood vaccination and COVID-19 vaccination for health workers and for adults [14]. We will adapt the BeSD COVID-19 vaccination tools for adults to our local context [31].

### 2.2. Study design

This research is designed as multiple case studies of four sites. Case study research is empirical research that explores a contemporary phenomenon within its real-world context [32]. Case studies are especially useful when the boundaries and interface between the “phenomenon” (or “the case”) and the “context” are not so clear. Case studies usually rely on multiple sources of data. In our study, the “phenomenon” is the introduction of COVID-19 vaccination into the four selected communities in South Africa with their different contextual factors, histories, and experience of COVID-19. Case studies are dominated by ‘how and why’ research questions. By adopting an approach that relies on more than one data collection method, it is possible through a case study to describe, explore, and explain the “how and why” of possible vaccine hesitancy without presuming at the outset to know the contextual and other factors that are shaping local behaviour. The research methods will be carefully replicated at each site; the underlying goal being either that the research will provide the replication of results across sites or that the case studies will provide theory that can predict either similar or contrasting findings across different communities [32]. In either case, multiple case studies will provide evidence that will be helpful to the activities of the South African Government in addressing vaccine hesitancy.

### 2.3. Study settings

Table 1 shows the site selection of the four case studies. Convenience and purposive sampling were used in the selection of sites. Convenience sampling ensured that some sites which are well known to the study team with established research and community networks to facilitate speedy data collection can be used. Purposive sampling will allow the team to include a mix of sites from different South African provinces badly affected by COVID-19 and to include communities that reflect formal and informal urban contexts as well as peri-urban and rural environments. Study sites were selected in KwaZulu Natal, Gauteng, and Western Cape provinces, which were the three provinces with highest numbers of COVID-19 cases in South Africa in early 2021 [33]. In KwaZulu Natal Province we selected an urban community (Wentworth) and a semi-rural one (Sweetwaters). In Gauteng Province we selected an urban informal community known as Alexandra. Finally, in the Western Cape Province, an urban middle-class community was selected. The selected sites have markedly different populations as shown in Table 1.

**Table 1.**
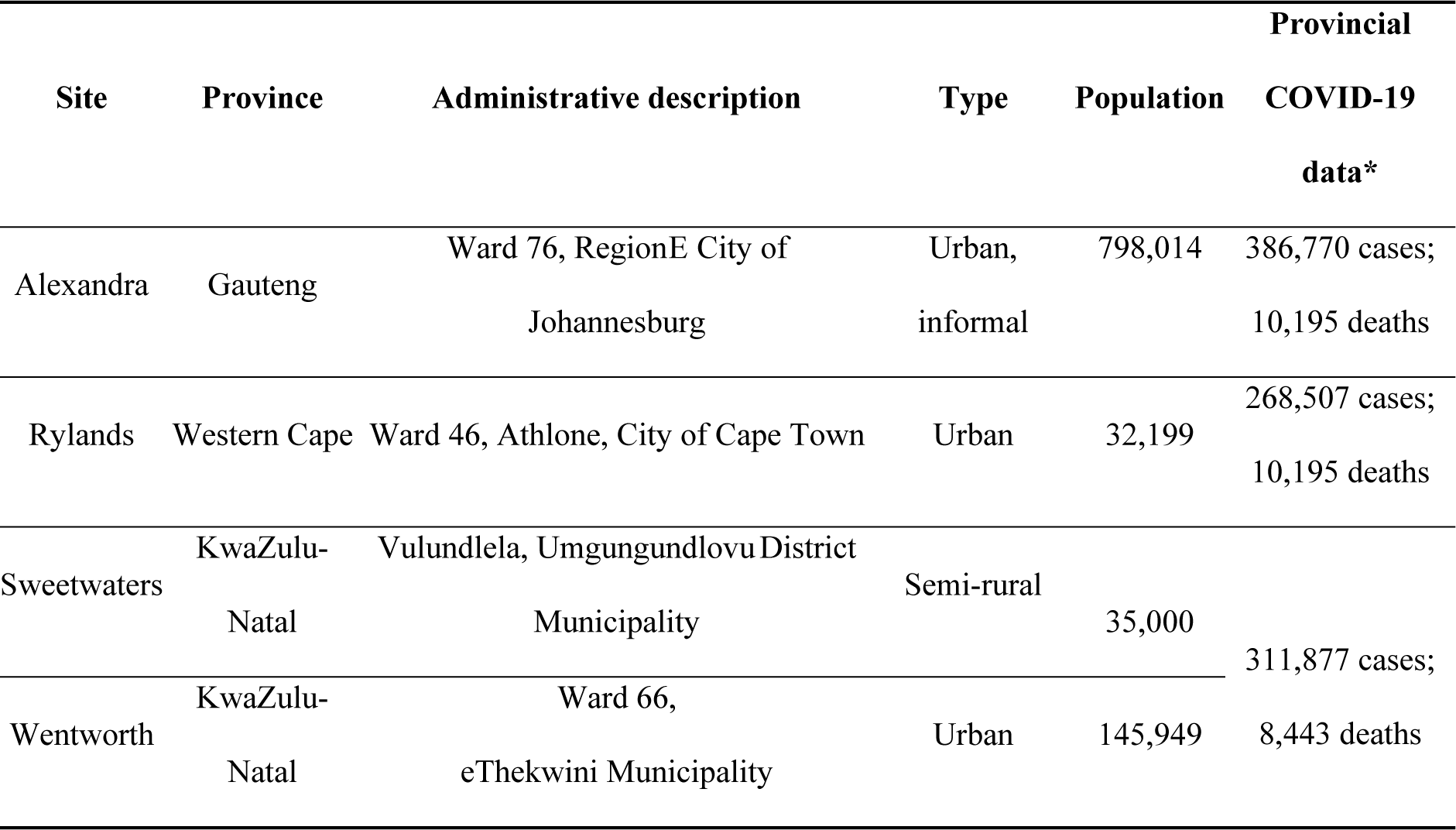
Characteristics of the four case study sites.

The source of the provincial COVID-19 data is the South African National Department of Health, as reported on 29 January 2021 [33].

### 2.4. Study phases

At each case study site, teams will use a mixed methods approach to data collection with quantitative and qualitative elements to address all research objectives. Figure 1 illustrates the overall approach to the study that is to be replicated in each of the case study sites. At each site data collection and the dissemination of findings will be organised into three distinctive phases.

**Figure 1.**
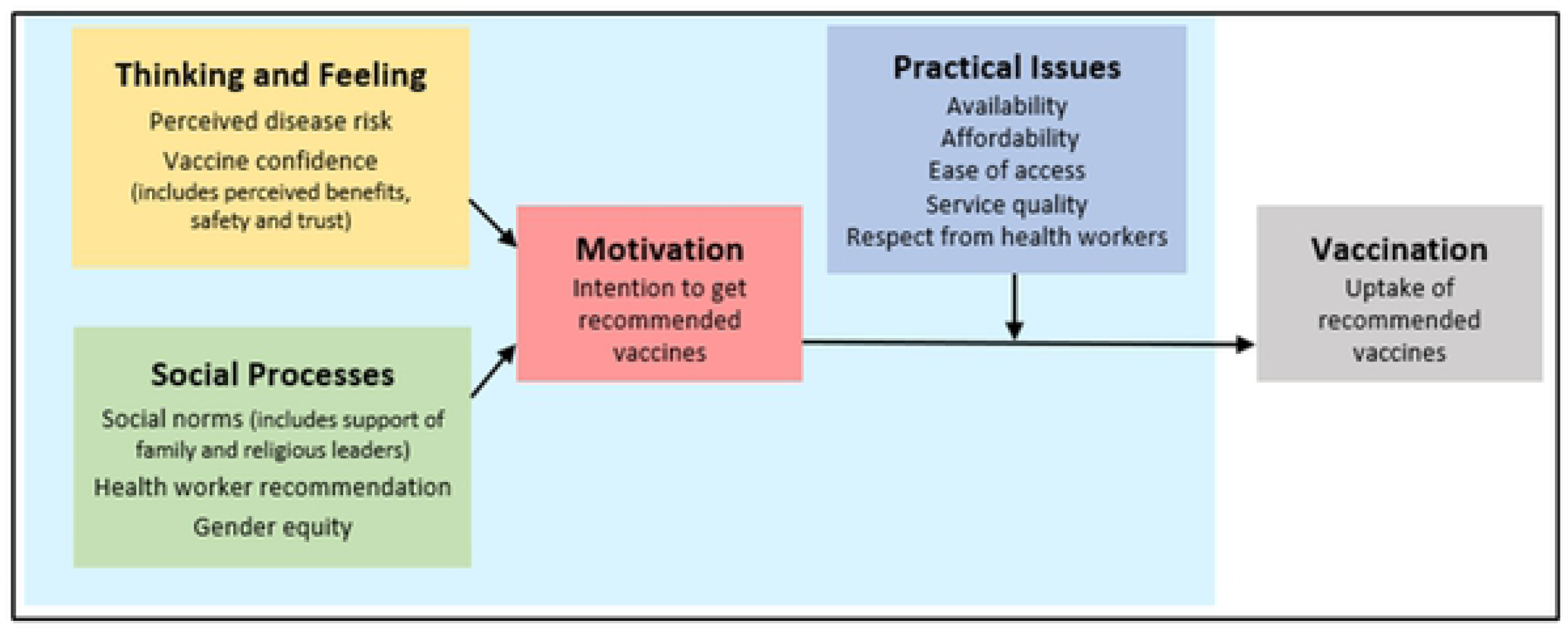
The Behavioural and Social Drivers of Vaccination Framework. Source: The BeSD Working Group [11, 14].

**Figure 2.**
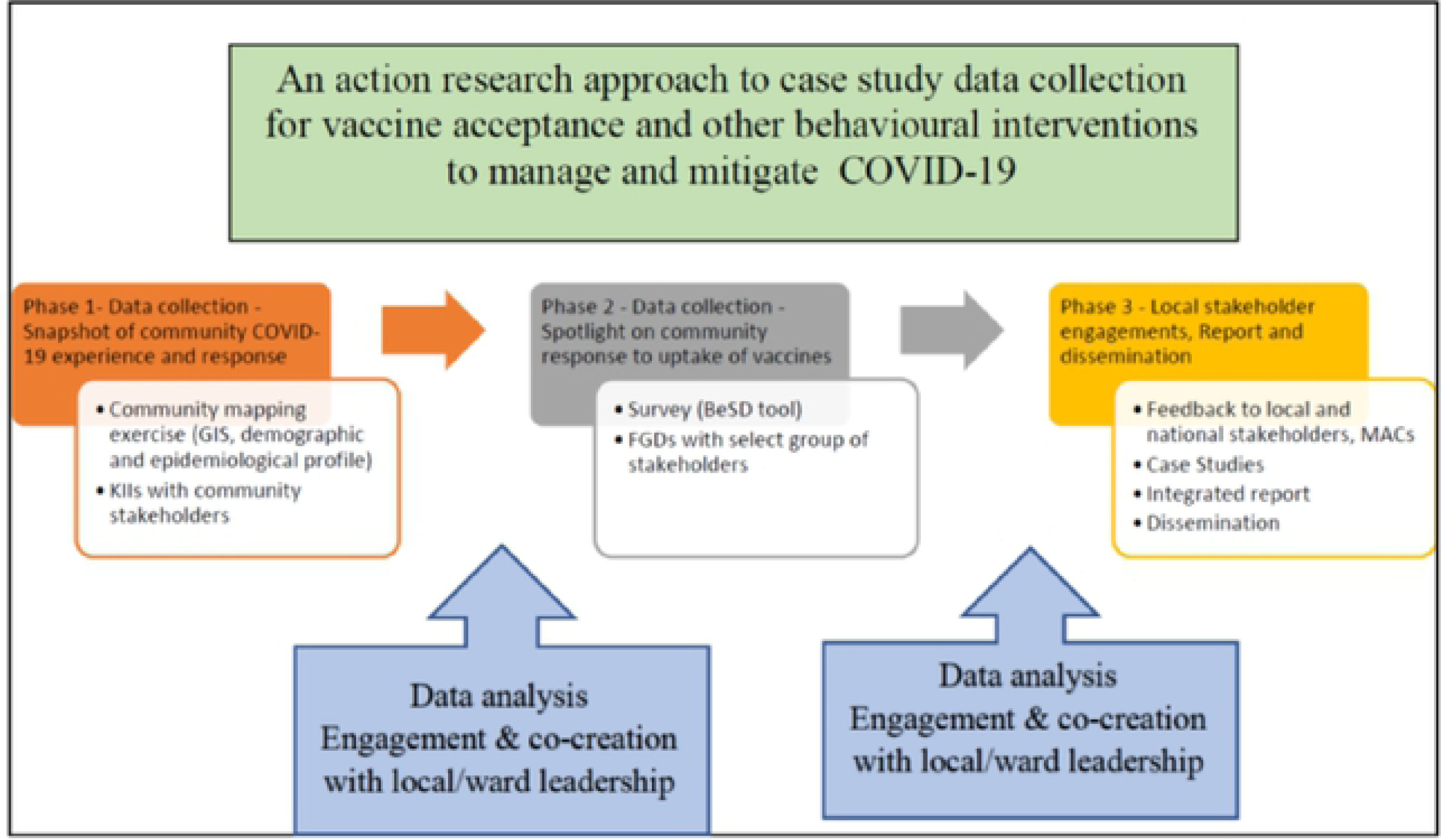
The three phases of data collection and dissemination for the VaxScenes Study. (BeSD, behavioural and social drivers of vaccination; KIIs, key informant interviews; FGDs, focus group discussions; GIS, geographic information system; MAC, ministerial advisory committees)

**Phase 1** will provide a preliminary snapshot of the community experiences of COVID-19 and the responses to it. This will be achieved using GIS maps to visualise the spatial arrangements for infrastructure, housing, employment, and public and private services, as well as population centres and COVID-19 statistics. Thereafter key informant interviews (KIIs)with community stakeholders and focus group discussions (FGDs) designed as expression sessions will provide a narrative about the COVID-19 experience in the local community. The local map and narrative will be shared with key informants at a workshop where the findings will provide the basis for the selection of population groups for a deeper investigation of vaccine hesitancy in phase 2.

In **Phase 2**, the case study research design will throw a spotlight on community acceptance of COVID-19 vaccines. A quantitative survey and further FGDs will be conducted with groups in the community. The findings of the survey and focus group discussions will be shared with the key informants at a second workshop and this will form the basis for stakeholders co-creating strategies to maximise vaccine acceptance within their ward.

**Phase 3** will involve cross case study syntheses as appropriate, and the presentation of findings to national role-players including the National Department of Health (NDoH), the Ministerial Advisory Committee (MAC) on COVID-19 Vaccines, the MAC on COVID-19, and the Behaviour Change Communication MAC. In Phase 3 researchers will prepare an integrated report across all four case study sites. Academic papers will be prepared and complemented by a comprehensive literature review of vaccine hesitancy and behavioural interventions as specifically experienced in Africa and in other cases where local perspectives about vaccine acceptability are integrated into national vaccine programmes.

### 2.5. Study population

Table 2 summarises the study population for both the qualitative and quantitative components. The study will only recruit people who are aged 18 years or older.

**Table 2.**
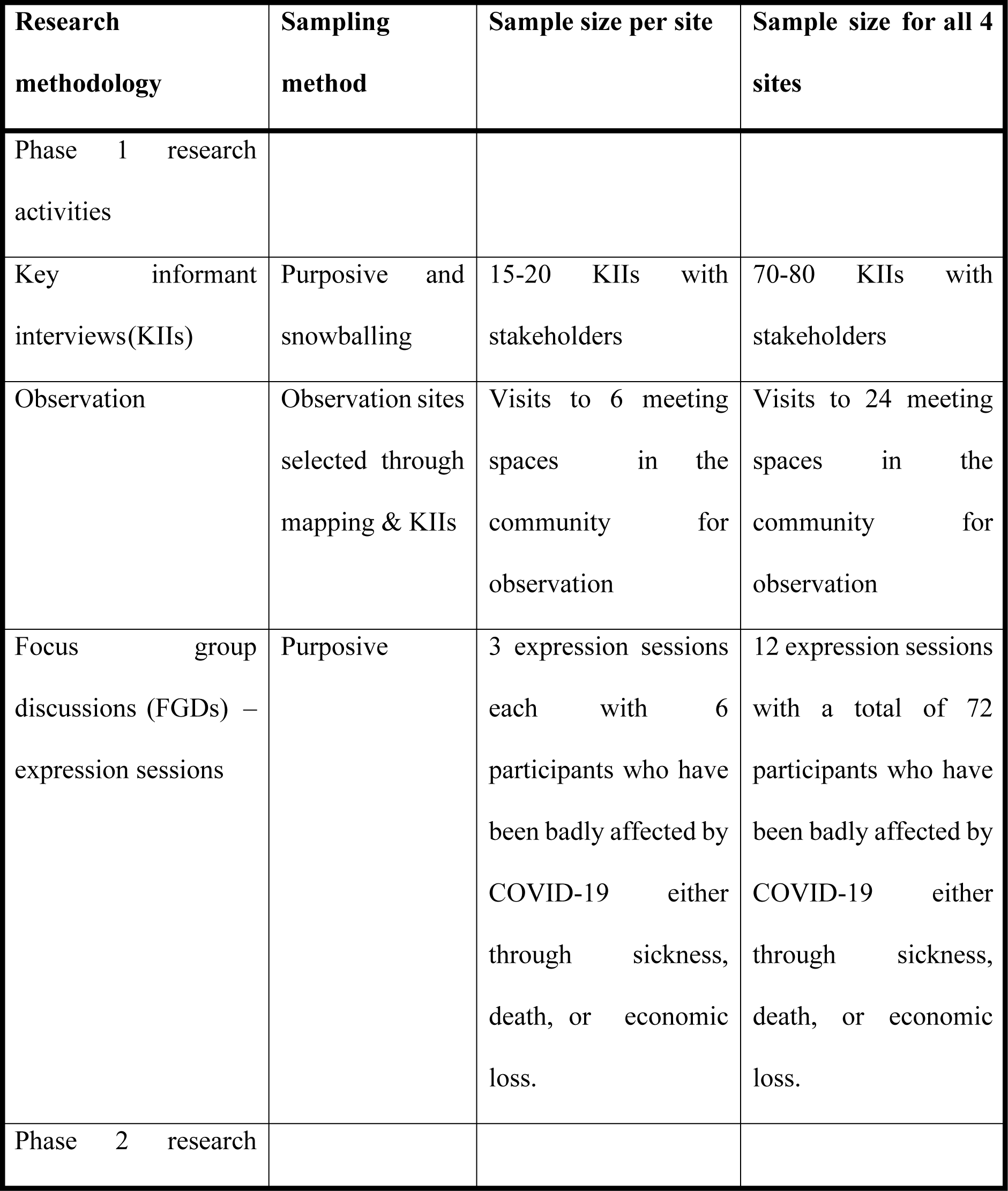
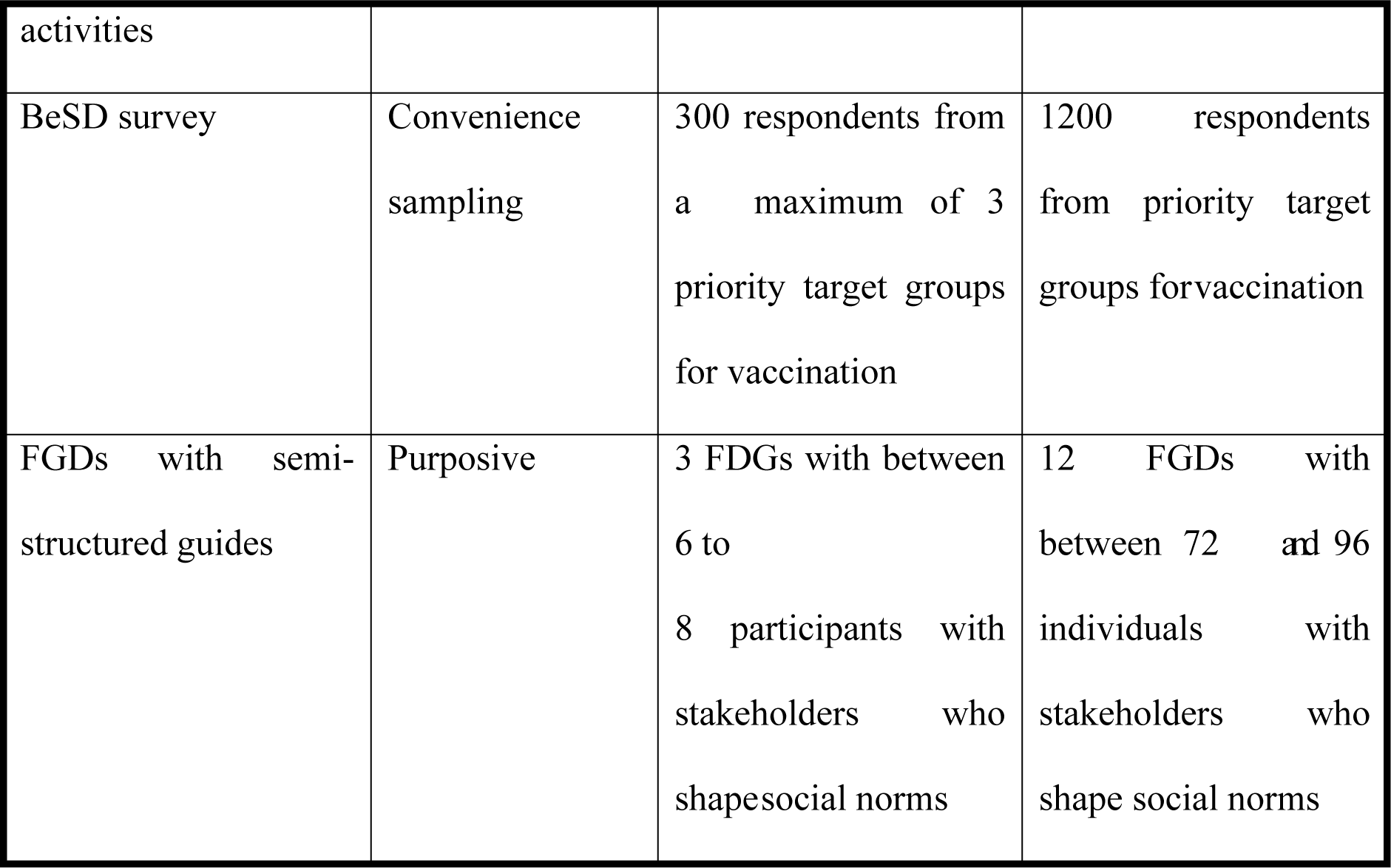
Sampling method and size per research methodology type at four case sites.

#### 2.5.1. Study population for qualitative component

During Phase 1 we will conduct 15-20 key informant interviews (KIIs) with stakeholders who are purposively selected. The proposed list of ward-based key informants includes but is not limited to local political and traditional leadership, clinic and health service leaders, school principals, faith-based leaders, business leaders, government officials, local taxi associations, local civic leaders, local trade union leadership, and private healthcare practitioners. It is expected that key informants at each site will include many of the individuals listed above, but also that at each site a unique list of key informants will emerge iteratively. During the interviewing of key stakeholders, researchers will enquire after the names of other potential key informants to ensure that influential leadership at the local level is not inadvertently overlooked during the mapping exercise. Snowball sampling would ensure that the research team identifies all significant individuals shaping the local COVID-19 response who may, for example, be outside of the immediate community such as provincial and/or national role-players.

Participants for both types of FGDs will be purposively identified. Three FGDs of each type will be held at each site. During Phase 1 no more than six participants will be recruited into the expression sessions. The sessions will be implemented in a group setting consisting of no more than six participants. The small sample size here is deliberate, to allow for meaningful engagement in the focus group discussions. Participants who have been badly affected by COVID-19 either through sickness, death, or economic loss will be recruited into the expression sessions. A social worker will be available at each site to ensure that any trauma experienced during or after the sessions can be appropriately referred. In Phase 2, between six and eight participants will be purposively recruited into the FGDs that are to be supported with a semi-structured interview guide. Specifically, this group of respondents are those best placed to influence social norms around vaccination in the community. The participants for these FGDs will be identified at the end of Phase 1 and in discussion with community stakeholders. This could include healthcare workers, faith-based leaders, community leadership, and other community and social workers.

#### 2.5.2. Study population for the quantitative component

The selection of survey respondents will be guided by priority populations in line with the rollout phases and target groups for COVID-19 vaccination as determined by the NDoH. The findings of Phase 1 research and consultation with local stakeholders will also determine the final selection of the survey respondents. It is expected that it may include priority groups at risk of COVID-19 (the elderly), essential service workers identified in each study site (for example, teachers, taxi drivers, and people working in institutional care facilities) and other adult men and women in the selected communities. The participants should also be resident or working in the community where the survey is to be conducted. The study will use non-probability convenience sampling because it is a cost- and time-effective method. It is proposed to collect 300 completed survey questionnaires at each site. Non-probability sampling can account for smaller samples such as 300 per site. A sample comprising at least 300 participants per site allows for the detection of small correlations (r=0.2) with at least 95% power [34].

### 2.6. Data collection

Three discrete but interlinking components of data collection will be undertaken for each case study site. These are: desk review of demographic and epidemiological profiles and available GIS mapping and literature; qualitative data collection methods; and quantitative data collection using a questionnaire.

#### 2.6.1. Desk based methods

A desktop mapping exercise using existing GIS and COVID-19 related document collection will complement qualitative and quantitative data collection at each site. The desk review will aim to create a comprehensive picture of each community. This will include a demographic and epidemiological profile, and mapping of the local community using general spatial planning data commonly stored on GIS and the collection of documents that describe the COVID-19 response in the community. A COVID-19 relevant picture of each community will be created, using GIS to support this, and will include: COVID-19 epidemiological profile, population (numbers, priority and at-risk groups, and population centres), services and local infrastructure (schools, clinics, transport routes) taxis, buses, private, civil society (faith-based, women’s groups, non-governmental organisations, government offices), relevant government departments, police, private sector (large employers and trade union representatives, private health care, COVID-19 response (who has done what where and the size of the spend). The desk-based mapping and evidence review will inform the sampling for the key informant interviews with stakeholders described later.

#### 2.6.2. Qualitative methods

##### 2.6.2.1. Key informant interviews

The mapping exercise will identify key stakeholders in each community to be approached for interview. Researchers will use a semi-structured interview guide (see Appendix A1), adapted from the BeSD qualitative tools for COVID-19 vaccination, for KIIs. The latter will be conducted face-to-face at the community level or over virtual platforms; depending on COVID-19 conditions at the time, participant preferences, and other practical considerations. The KIIs will explore a range of themes including: participants’ personal, family and community experiences of COVID-19; the impact of COVID-19 on personal and work life; experience of varying lockdown regulations; experience of adherence to NPIs; understanding of COVID-19 vaccines; how COVID-19 has shaped theirleadership role in the community; and expectations for the future. All research teams will consist of a mix of local language and English-speaking researchers. With permission of participants, interviews will be audio-recorded and thereafter translated as necessary and transcribed.

##### 2.6.2.2. Observation

On-site researchers will conduct observations of community life in the case study sites. These observations will be centred around locations where community members meet, such as shopping centres, street trading spaces, and/or taxi ranks. The purpose of the observations will be to better understand local community life as well as to document observation about NPI adherence as well as COVID-19 related media and facilities; such as for hand washing.

##### 2.6.2.3. Focus group discussions

Two types of FGDs will be conducted. The first type will apply an approach called “expression sessions” [35]. The second type of FDG will rely on a semi-structured FGD guides. Expression sessions will be conducted in Phase 1 of the research activities. The second type of FGD will follow in Phase 2. All FGDs will be conducted face-to-face at the research site and will follow COVID-19 precautions including physical distancing, the wearing of face masks, and a well-ventilated venue. FGDs will be capped at six participants for expression sessions and eight for other FGDs. The tools will be piloted in one community and amendments made thereafter. All FGDs will be audio recorded with the permission of participants. Thereafter recordings will be translated as necessary and transcribed.

###### 2.6.2.3.1. Expression session focus group discussions

Expression sessions will be facilitated with participants in the community who have been severely affected by COVID-19 through personal ill-health, family loss, and/or economic adversity. An expression session is a newly developed approach to collect verbal, visual, and audio-visual data from participants [36]. This approach is informed by the photovoice methodology where participants are required to take photos in response to research questions [37]. The approach encourages active participation in research, and meaningful reflection and engagement with research questions, which is important in the context of COVID-19 and vaccine hesitancy. Expression sessions encourage participants to respond to probes by sharing ‘something’ rather than photos only. ‘Something’ could be images, photos, videos (self-taken, downloaded online, or from a book or magazine), a song (audio or lyrics), a drawing (self-drawn or a picture of a drawing), or a poem (self-written or not). Participants will be encouraged to bring their ‘somethings’ to a FGD. Participants will receive a brief formative training session on the methodology along with a set of two research probes to which they can respond by bringing ‘something’ (see Appendix A2). Following the formative session, participants will have one week to gather their ‘somethings’. Expression session FGDs will be facilitated by two researchers and will be conducted in the dominant or preferred language of the group. Each team will include a researcher with local language capacity. Participant information sheets and consent forms will be translated into the local language.

###### 2.6.2.3.2. Focus group discussions with semi-structured guides

Conventional FGDs will be conducted with three stakeholder groups who are able to address vaccine hesitancy in the community and provide support to the local response to improve acceptance and uptake of vaccines. These FDGs will focus on how to strengthen the community response to vaccine hesitancy and to promote on-going COVID health seeking behaviours. A semi-structured FGD protocol is found in Appendix A3. FGDs will be facilitated by two researchers. It is anticipated that most of these FGDs can be conducted in English but researchers with local language capacity will be included in each research team. Participant information sheets and consent forms will be translated into the local language.

#### 2.6.3. Quantitative methods

For the quantitative component of the study, we adapted the BeSD quantitative COVID-19 vaccination tool for adults [14]. The tool consists of a questionnaire that assesses four domains i.e., what adults think and feel about COVID-19 vaccines; social processes that drive or inhibit COVID-19 vaccination; individual motivations (or hesitancy) to seek COVID-19 vaccination; and practical factors that shape the experience of seeking and receiving COVID-19 vaccination [14]. The BeSD quantitative tool adapted for this study is provided in Appendix A4. The survey will be conducted in English, Afrikaans, and isiZulu since these are predominant languages spoken in the selected communities. Participant information sheets and consent forms will also be translated into the local language. The survey tool will be piloted among 100 adults in one of the first community sites. Where needed additions will be made to the survey to assist with the translation of specific terms. Data will be captured onto tablets during face-to-face interviews conducted by well-trained researchers working in each community site. The researchers will wear masks, practice physical distancing when approaching potential participants, use hand sanitisers regularly, and adhere to all other COVID-19 protocols. Should the period for interviews in any given community coincide with the period of lockdown, face-to-face interviews will be replaced with online questionnaires.

#### 2.6.4. COVID-19 precautions

The research team will adopt an approach to data collection that is compliant with all the rules pertaining to the COVID-19 response. It is expected that some qualitative data collection will be virtual and meetings to discuss findings with local stakeholders may be virtual where appropriate. Onsite researchers will use personal protective equipment (including face masks and hand sanitisers) and with provide PPE to FGD participants. Meetings and FGDs will be made in well ventilated venues and seating will be appropriately physically distanced.

### 2.7. Data analyses

Iterative analyses of all data will occur at each community site as the research team moves from Phase 1 to Phase 3. Data will be triangulated across methods to promote validity. Case-specific and cross-case syntheses will be undertaken as appropriate and as determined by the findings.

#### 2.7.1. Qualitative data analyses

Qualitative data will be audio recorded, transcribed, and translated where required. Transcripts will be organised and stored using qualitative data software such as MaxQDA and ATLAS.ti (version 9) to facilitate coding and analyses of data. Qualitative data will be subject to thematic analysis [38]. A code book will be developed for the data including both inductive and deductive codes [39], but emphasis will be placed on allowing the data to determine codes [40]. The codes will be discussed between the four qualitative researchers involved in the case studies and with the four study principal investigators. An inter-coder agreement will be established with a portion of the data to verify and enhance data credibility.

#### 2.7.2. Quantitative data analyses

Quantitative data will be cleaned and stored in the Statistical Package for Social Sciences software, version 27.0 (IBM SPSS Inc, Chicago, IL). Thereafter both the SPSS version 27.0 and R/Rstudio v3 softwares will be used for data analyses. The survey data will be summarised as counts and percentages for categorical variables and means with their standard deviations for continuous variables. Analysis of the variance (ANOVA), chi-square tests, and equivalents will be used as appropriate for group comparisons. Logistic regression models will also be used to evaluate the association between selected characteristics and vaccine hesitancy as well as adherence to NPIs. A basic model will be adjusted for age and sex. Expanded multivariable models for the outcome “vaccine hesitancy” will be further adjusted for significant predictors in basic models. Additional data analyses will include subgroup analyses conducted using variables such as race, sex, socio-economic status, geographical location, education levels, and occupation to match areas specific needs. A P-value less than 0.05 will be used to indicate statistically significant results.

### 2.8. Ethics

This study proposal was approved by the Human Research Ethics Committees of the University of the Witwatersrand in Johannesburg, South Africa (reference: H21/02/05) on 25 March 2021; the Human Sciences Research Council of South Africa (reference: REC 12/04/21) on 20 April 2021; and the South African Medical Research Council (reference: EC022-5/2021) on 27 May 2021.

Every effort will be made to ensure that the research process is socially sensitive and respects the rights of all participants. Information sheets will be provided to all potential participants (as hard or electronical copies as the case may be) as well as verbal briefing on the nature and purpose of the interviews or FGDs, emphasising the right of individuals to refuse or withdraw from the interview or FGD at any point. A consent form for participation and for audio-recording will be provided. All participation is voluntary. Participants at face-to-face FGDs will be provided with refreshments.

Personal identifiers during KIIs and FGDs will not be used in the reporting and all data will be reported in a summarised form (sex, age) without attributing comments or information to individuals. FGD participants will be informed that while all steps will be taken towards maintaining confidentiality and anonymity, this cannot be guaranteed due to the nature of sharing encouraged by FGDs. While FGD participants will know each other’s identity and the information shared, an environment of private sharing will be encouraged; highlighting the importance of maintaining confidentiality for all participants. Furthermore, a local social worker will be available at each site to support or refer any participant that experiences any psychosocial concerns during or after the expression session FGDs. All data will only be used for research purposes and kept with utmost confidentiality, and they will not be accessed by anyone else but the research team.

Respondents will be reimbursed for their participation in the study. This is ethically sound and is aligned to the “time, inconvenience, and expenses” (TIE) model [41] and is endorsed by the National Health Research Ethics Council of South Africa [42]. The reimbursement amounts proposed for the study are aligned to TIE that reimburses participants for their time at a rate on par with unskilled labour rates, for their inconvenience and for any expenses (e.g., transport, childcare). The reimbursement for KIIs and FGDs is 150 South African Rands (ZAR) and for the survey is ZAR 100.

### 2.9. Study team

The study team is multidisciplinary, consisting of specialists in public health, epidemiology, social science, vaccinology, psychology, political science, project management, and health promotion. The team is led by four principal investigators from the three South African institutions, namely, the Human Sciences Research Council (H.v.R), the Sarraounia Public Health Trust (N.C. and N.P.), and the South African Medical Research Council (C.S.W.).

## 3. Discussion

Previous studies have shown high levels of vaccine hesitancy in South Africa [9, 13, 43]. These high levels of are most likely to be driven by multiple factors including personal, interpersonal, community, and social. We therefore propose in this study to determine and co-create with community stakeholders a comprehensive understanding of vaccine hesitancy and opportunities to support the promotion of COVID-19 health seeking behaviours in South Africa. The conceptual framework for the study is based on the socio-ecological model of behaviour change [29] and the BeSD framework for COVID-19 vaccination [14].

Existing vaccine hesitancy measurement tools were mostly designed for use in and validated in high-income countries [10, 15]. Given that vaccine hesitancy is context specific, there is a need to adapt and validate existing COVID-19 vaccine hesitancy tools for use at national and community levels in South Africa. The BeSD tools, which were recently developed under the auspices of WHO, measure both behavioural and social drivers of vaccination. At the time this study was conceived in early 2021, the BeSD model of vaccine hesitancy had not been validated in an African setting. The proposed study will therefore provide insights into the applicability of a novel vaccine hesitancy model in Africa.

The backdrop to the proposed community-based case studies is that the benefits of models of behaviour change that are focused purely on knowledge deficit are limited. Understanding both the behavioural (individual) and social drivers of vaccine hesitancy is critical to increasing COVID-19 vaccine uptake in South Africa. The study applies the socio-ecological model to the investigation of vaccine hesitancy. This model considers the complex interplay between individual, relationship (interpersonal), community, and social factors. It allows us to explore the range of factors that render people conflicted about or opposed to COVID-19 vaccination. Although all levels of the socio-ecological model are relevant for communities, researchers and practitioners in health promotion are often poor at articulating the integration of these levels and how this determines effective health promotion strategies. One of the most effective applications of the socio-ecological model is at the community level. This is because the enablers or barriers to behaviour change in the four levels (i.e., personal, interpersonal, community, social) of the model can be readily identified. For example, if a local faith leader is speaking out against vaccination, it will most likely have an impact on the social norms of the local community. Similarly, if the local clinic has a reputation for providing poor services, then this too will affect vaccine uptake because it is a barrier to establishing an enabling environment. A bottom-up approach to understanding vaccine hesitancy provides the best opportunity to understand those factors that encourage or impede vaccine uptake. This is enhanced when community stakeholders are actively engaged in providing an interpretation of the intersection of the range of factors at the community level.

This mixed methods study is designed as an action research study that is responsive to the local context and is conducted to influence the ongoing management and mitigation of COVID-19 in South Africa. To address the issue of COVID-19 vaccine uptake, an integrated analysis is necessary to provide community stakeholders with a comprehensive understanding of the factors that will assist the identification of helpful health promotion activities at the local level to support COVID-19 vaccine rollout. Convenience sampling is best suited for this type of study, although this would limit the generalisability of the findings to all communities in South Africa. However, we believe that the study will make a substantial contribution to knowledge on COVID-19 response in South Africa, as the case study research will be conducted in four commonplace settings in the country.

## 4. Conclusions

The COVID-19 pandemic continues to have significant health, human, social, and economic impacts on South African society. COVID-19 vaccines and non-pharmaceutical interventions (NPIs) currently offer the most promising means to manage the pandemic. However, their success depends on high levels of uptake and adherence. The aim of this study is to determine and co-create with local stakeholders a comprehensive understanding of vaccine hesitancy and opportunities to support the promotion of other COVID-19 health-promoting behaviours. The study will utilise a mixed-methods, multiple case study design, informed by the socio-ecological model of behaviour change. The study will provide ground-up, locally responsive, and timeous evidence on the factors influencing COVID-19 vaccine acceptance and other health-seeking behaviours to inform the management and mitigation of the pandemic in South Africa. It will also provide insights into the applicability of various global vaccine hesitancy models and research tools to a middle-income country in Africa.

## Data Availability

No datasets were generated or analysed during the current study. All relevant data from this study will be made available upon study completion.

## Author Contributions

Conceptualization, C.S.W., N.C., N.P., H.v.R; methodology, C.S.W., N.C., N.P., S.C., C.G., PDK, Z.E., S.P., H.v.R; writing—original draft preparation, C.S.W., N.C., N.P., H.v.R.; writing—review and editing, S.C., C.G., Z.E., S.P., G.H., J.S., A.J. and PDK; project administration, N.C., N.P., C.G., Z.E., G.H., J.S; and funding acquisition, H.v.R. All authors have read and agreed to the published version of the manuscript.

## Informed Consent Statement

Each participant will provide written or digital informed consent.

## Acknowledgments

This study is conducted by The VaxScenes Study Team which includes Heidi van Rooyen (Human Sciences Research Council (HSRC)), Charles Shey Wiysonge (South African Medical Research Council (SAMRC)) Nancy Coulson (Sarraounia Public Health Trust), Nirvana Pillay (Sarraounia Public Health Trust), Saahier Parker (HSRC), Gregory Houston (HSRC), Marilyn Couch (HSRC), Candice Groenewald (HSRC), Zaynab Essack (HSRC), Londiwe Deborah Shandu (HSRC), Nonkululeko Khuzwayo (HSRC), Nobukhosi Ncube (Sarraounia Public Health Trust), Jane Simmonds (SAMRC), Sara Cooper (SAMRC), Anelisa Jaca (SAMRC), Phelele Bhengu (University of Cape Town), Edison Mavundza (SAMRC), Patrick Katoto (SAMRC).

